# In vivo kinetics of SARS-CoV-2 infection and its relationship with a person’s infectiousness

**DOI:** 10.1101/2021.06.26.21259581

**Authors:** Ruian Ke, Carolin Zitzmann, David D. Ho, Ruy M. Ribeiro, Alan S. Perelson

**Affiliations:** Theoretical Biology and Biophysics Group, Theoretical Division, Los Alamos National Laboratory, Los Alamos, NM 87545, USA; New Mexico Consortium, 4200 West Jemez Road, Los Alamos, NM 87544; Aaron Diamond AIDS Research Center, Columbia University Vagelos College of Physicians and Surgeons, New York, NY 10032

**Keywords:** SARS-CoV-2, viral kinetics, viral transmission

## Abstract

The within-host viral kinetics of SARS-CoV-2 infection and how they relate to a person’s infectiousness are not well understood. This limits our ability to quantify the impact of interventions on viral transmission. Here, we develop data-driven viral dynamic models of SARS-CoV-2 infection and estimate key within-host parameters such as the infected cell half-life and the within-host reproductive number. We then develop a model linking VL to infectiousness, showing that a person’s infectiousness increases sub-linearly with VL. We show that the logarithm of the VL in the upper respiratory tract (URT) is a better surrogate of infectiousness than the VL itself. Using data on VL and the predicted infectiousness, we further incorporated data on antigen and reverse transcription polymerase chain reaction (RT-PCR) tests and compared their usefulness in detecting infection and preventing transmission. We found that RT-PCR tests perform better than antigen tests assuming equal testing frequency; however, more frequent antigen testing may perform equally well with RT-PCR tests at a lower cost, but with many more false-negative tests. Overall, our models provide a quantitative framework for inferring the impact of therapeutics and vaccines that lower VL on the infectiousness of individuals and for evaluating rapid testing strategies.

**Significance:** Quantifying the kinetics of SARS-CoV-2 infection and individual infectiousness is key to quantitatively understanding SARS-CoV-2 transmission and evaluating intervention strategies. Here we developed data-driven within-host models of SARS-CoV-2 infection and by fitting them to clinical data we estimated key within-host viral dynamic parameters. We also developed a mechanistic model for viral transmission and show that the logarithm of the viral load in the upper respiratory tract serves an appropriate surrogate for a person’s infectiousness. Using data on how viral load changes during infection, we further evaluated the effectiveness of PCR and antigen-based testing strategies for averting transmission and identifying infected individuals.

## Introduction

SARS-CoV-2 is a new human pathogen that causes COVID-19 (1). It is highly contagious and spread rapidly across the globe and has caused more than 3.8 million deaths worldwide as of June 2021. Extensive efforts to develop effective treatments and vaccines are underway, with some successful vaccines already in use (2, 3). At the molecular level, SARS-CoV-2 enters host cells via the cell surface receptor ACE-2 (angiotensin converting enzyme 2) (4). Structural analysis suggests that SARS-CoV-2 binds to the receptor >10-fold more efficiently than SARS-CoV-1 (5), partially explaining the comparatively high contagiousness of the virus (6-8).

It is likely that the ability of the virus to effectively infect cells in the upper respiratory tract (URT) allows the virus to reach a high viral load (VL) and be effectively transmitted even before symptom onset (9, 10). However, it is not clear how VL, infectiousness and symptom onset are quantitatively related. Previously, both VL and log_10_ VL have been used as surrogates for infectiousness of influenza (11) and SARS-CoV-2 (12, 13). A quantitative understanding of the relationship is critical for both non-pharmaceutical and pharmaceutical interventions. First, it would allow for more precise predictions of the infectiousness of infected individuals, including children and asymptomatic individuals, based on their VL measurements (14, 15). This would in turn help to better inform public health policy decisions. Second, as administration of vaccines or effective therapeutics should lead to lowered VLs (16), a quantitative understanding will inform how these changes impact infectiousness and, in turn, SARS-CoV-2 transmission (17). This becomes particularly important in current outbreaks such as that in India where increases vaccination has been proposed as one means of controlling the epidemic. Third, it would provide better insight into a person’s infectiousness throughout the course of infection and thus inform test strategies for work/school reopening, travel, etc. The effectiveness of test, trace and quarantine as control strategies heavily depends on the sensitivity and specificity of the tests and rate of testing being implemented (18). It was recently proposed that antigen tests with low sensitivity are preferred over highly sensitive RT-PCR tests, because of their potential for wide coverage and short turn-around time (12). However, the effectiveness of this strategy has not been evaluated based on VL and infectiousness dynamics inferred from data.

Here, we construct viral dynamic models of SARS-CoV-2 upper respiratory tract (URT) infection and a model linking VL to infectiousness. Mathematical modeling has been applied, by us and others, to understand SARS-CoV-2 infection in hospitalized patients and the potential impact of therapy (19-22). However, there were large uncertainties in model parameter estimates because in almost all studies, viral dynamic models were fit to data that were taken days after symptom onset without knowledge of the patients’ infection dates. We resolve this issue by using two unique sets of data to quantify key within-host parameters, and by using a variety of clinical and epidemiological data to inform the quantitative relationship between VL and infectiousness. Using this relationship, we further evaluate the effectiveness of testing strategies using either antigen or RT-PCR tests at different testing frequencies.

## Results

### Datasets

We collected two unique sets of URT VL data for model inference. The first, the “German dataset”, contains VL measurements from 9 individuals in the first cluster of infections in Germany (23). All individuals had mild symptoms. VLs were measured longitudinally starting several days after symptom onset. We excluded one individual (Patient 16 in Ref. (24)), because their first VL measurement was too long after infection. A unique feature of this dataset is that the detailed transmission history, including the infection dates and dates of symptom onset, were reported (24). However, this data does not have good sampling during the initial expansion before the peak VL. Thus, we include a second data set, the NBA (National Basketball Association) dataset, which was taken from a study where individuals (staff and players) were regularly tested during an NBA tournament in 2020 (25). We selected 9 individuals sampled frequently, including during the virus expansion phase. Below, we show that these unique features of the two datasets allow us to jointly infer the within-host SARS-CoV-2 dynamics.

### Dynamics of early infection

The SARS-CoV-2 dynamics in the URT are typical of an acute respiratory infection, i.e., VLs increase to a viral peak and decline afterwards (Fig. 1). Thus, we constructed a target cell limited (TCL) model and an innate response model using frameworks developed for influenza infection (26, 27) (see Methods and *SI Appendix*). In particular, in the innate response model, we assumed that innate immune mediators, such as interferons, put target cells into an antiviral state that is refractory to viral infection (27). We first fit these two models to the NBA dataset to estimate the time of infection. Because multiple measurements were taken before peak VL in the 9 individuals we chose to study, the times of infection can be estimated relatively reliably. Both the TCL and the innate response model gave similar estimates of infection time (Table S1).

**Figure 1.**
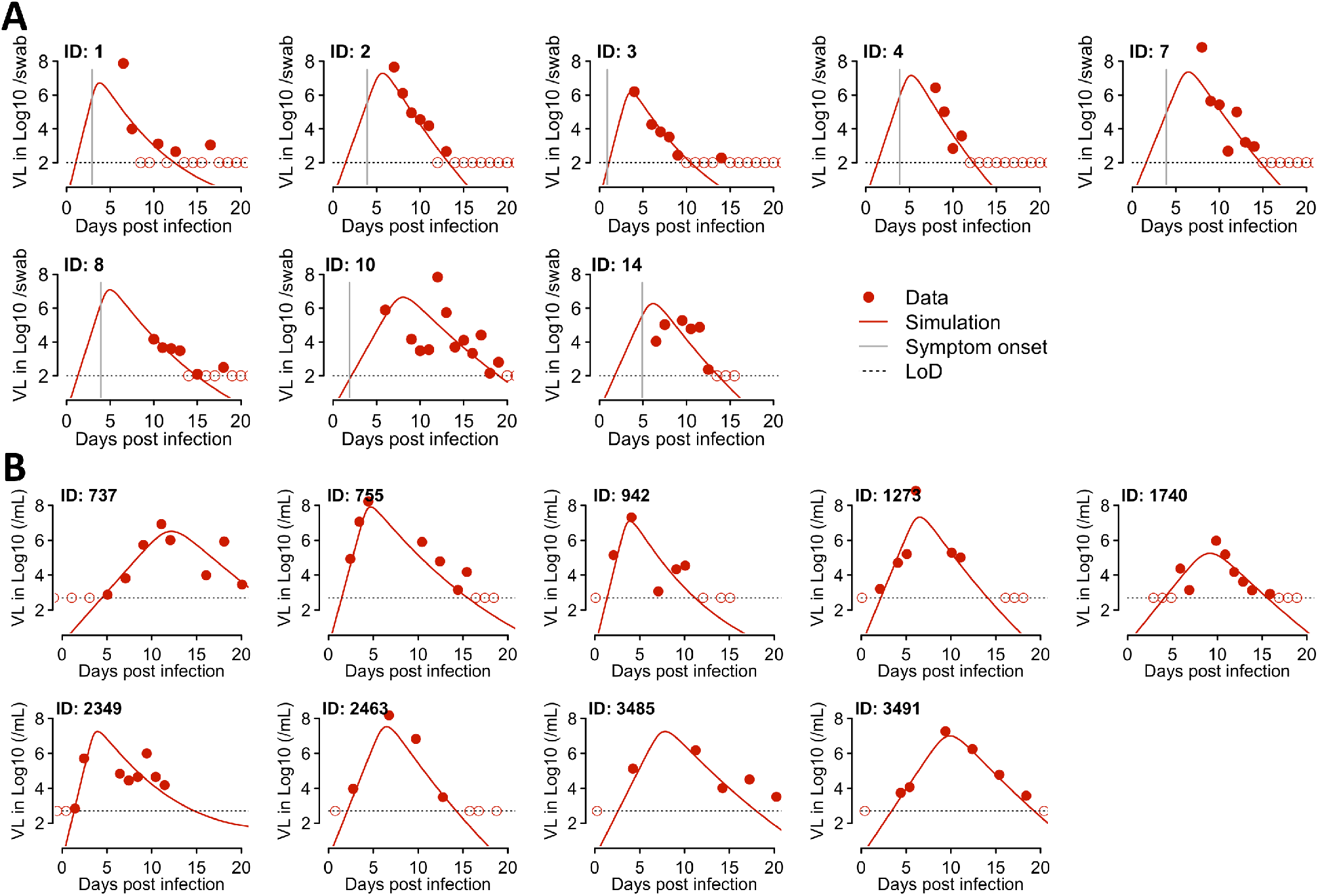
Fitting results of the innate response model to the VL data from two studies. **(A)** Fitting results to data from 8 individuals in the Germany study, i.e., Wolfel et al. (23). The model (solid lines) was simulated using the best-fit individual parameter values estimated by a non-linear mixed effect modeling approach (Table 1 and 2). Symbols (red dots and circles) show the data from pharyngeal swabs. Circles indicate data points below the limit of detection. Vertical grey lines denote the time of symptom onset as reported in Ref. (24). Horizontal dashed black lines show the limit of detection (LoD). **(B)** Fitting results to data from 9 individuals in the NBA study as reported in Kissler et al. (25) with symbols and colors as in (A).

We then fit the TCL model and the innate response model to the data from both datasets simultaneously using a nonlinear mixed effect modeling approach (see Methods). We also tested variants of the innate response model that assume the innate immune mediators block infection of target cells or reduce virus production from infected cells (see *SI Appendix*). According to the Akaike information criterion (AIC) scores, the best model overall is the original model assuming the innate immune mediators convert target cells into refractory cells (Table S2). This model fits both datasets well (Fig. 1) and it describes both the upslope and downslope of the viral dynamics in the NBA dataset. This gives confidence in our model predictions of the early viral dynamics for individuals in the German dataset. We then tested if there is any difference in estimated parameter values between the two datasets by including the source of the dataset, i.e., the NBA or the German dataset, as a covariate in the model fitting. We found that there was no statistical support for including the origin of the datasets as a covariate (Table S2). Therefore, we use the innate response model without the covariate for further analysis and term this model the innate response model for short.

According to the best-fit parameter values, the infected cell death rate *δ* is 1.7 d^-1^ on average (Table 1). Because the model includes an eclipse phase of length 1/*k*, where *k*=4 d^-1^, the average lifespan of infected cells is 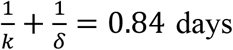. We estimated that the within-host basic reproductive number, R_0,within_, varies over a range between 2.6 and 14.9, with mean 7.4 (SD: ±3.8) (Table S3).

**Table 1.** Estimated population parameter values from the best model, i.e., the innate response model. The means and standard deviations are derived assuming that individual parameters follow log-normal distributions.

| Parameter | Description | Mean (population estimate) | Standard deviation |
| --- | --- | --- | --- |
| $\beta$ | Infectivity parameter constant | $3.2 \times 10^{-8}$ ml/RNA copy/day | 0.50 |
| $\delta$ | Death rate of infected cells | 1.7 /day | 0.23 |
| $\pi$ | Composite parameter for virus production and sampling | 45.3 /ml/day | 0.24 |
| $\Phi$ | Rate constant for the interferon-induced conversion of target cells to refractory cells | $1.3 \times 10^{-6}$ /cell /day | 1.95 |
| $\rho$ | Rate at which refractory cells become target cells again | 0.0044 /day | 0.20 |

We further tested how robust our estimates are with respect to variations in the fixed parameter values in the model by varying each of those in the ranges shown in Table 2 and then re-fitting the model to the data. Across the scenarios examined, the estimates of the death rate of infected cells were very consistent between 1.6 and 1.9 d^-1^ and the mean R_0_,within ranged between 5.8-8.9 (Table S4). Overall, the estimated parameters and viral dynamic characteristics were robust against variations in the fixed parameters (Tables S4).

**Table 2.** The fixed parameters in the viral dynamic models and their values.

| Parameter | Description | Values | Values tested in sensitivity analyses |
| --- | --- | --- | --- |
| $T_0$ | Total number of (infection free) target cells | $8 \times 10^7$ cells | NA |
| $E_0$ | Initial number of infected cells | 1 cell | 5, 10 cells |
| c | Virus clearance rate | 10/day | 5 and 20/day |
| k | 1/the eclipse phase duration | 4/day | 3 and 6/day |

### Probability of transmission

We next examine how VL is related to the infectiousness of a person by constructing a probabilistic model to describe the various steps in viral transmission from viral shedding to establishment of infection (see Fig. 2A for a schematic). We define infectiousness as the probability that an infected person, i.e., a donor, will shed one or more infectious viral particles leading to successful infection of a recipient for a typical contact of relatively short duration, 𝜏. The typical contact here is defined the same as in the epidemiological survey study in Mossong *et al*. (28). Note that the probability defined here only characterizes the infectiousness of a person arising from virus dynamics in the URT given a contact, and it does not assume any frequency of typical contacts. The expected number of transmissions that a person causes can be calculated if the contact pattern of the person is known.

**Figure 2.**
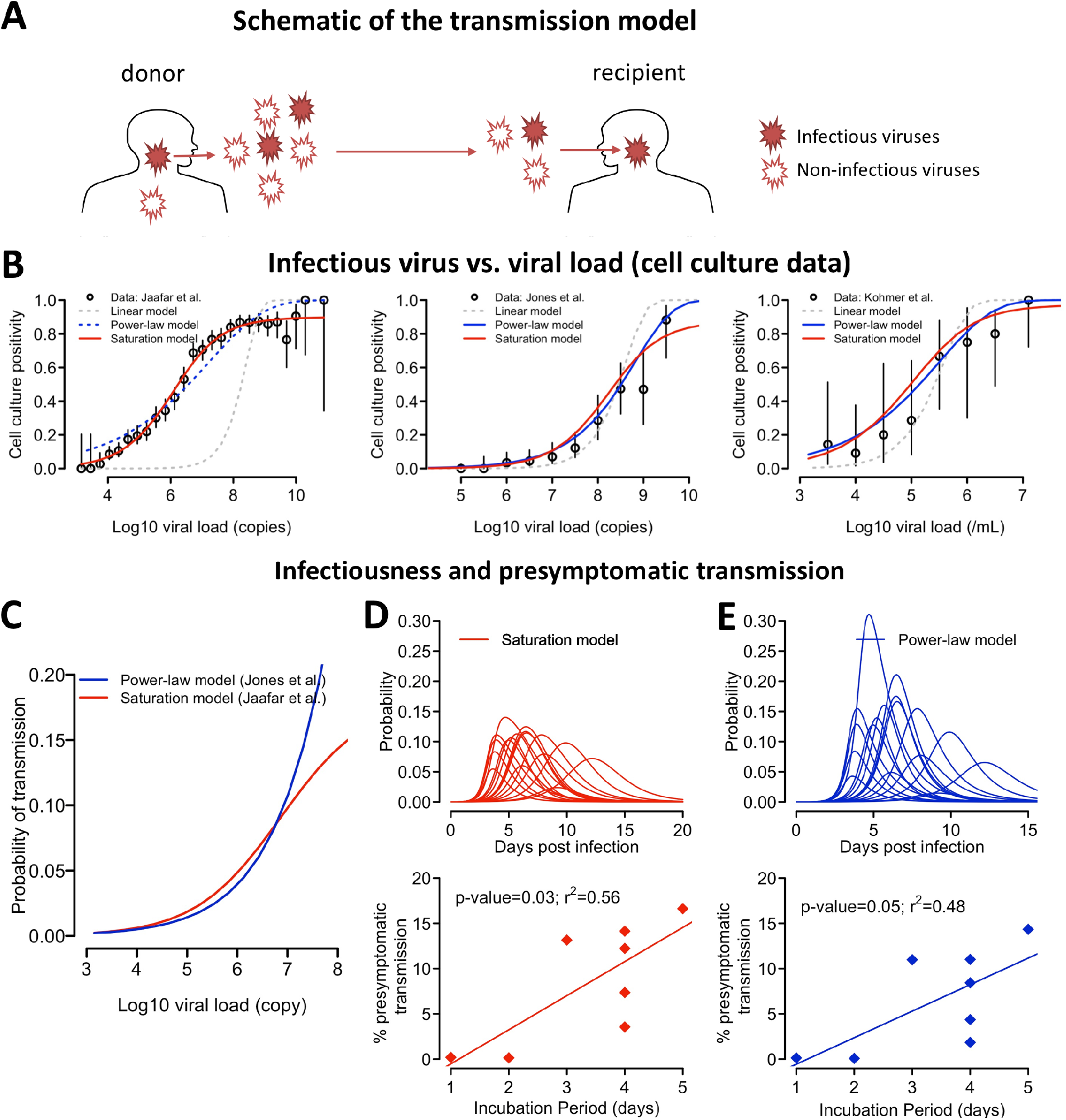
The relationship between VL and host infectiousness. **(A)** A schematic of the probabilistic model describing the steps in a transmission event. A donor sheds both infectious and non-infectious viruses, of which some infectious viruses may reach a recipient during a close contact and establish an infection. **(B)** Best-fit of the three models, i.e. the linear model (grey), the power-law model (blue) and the saturation model (red), to the data from Jaafar et al. (29), Jones et al. (30) and Kohmer et al. (31). Open circles denote the percentage of cell culture positivity reported, and vertical lines denote the 95% confidence intervals calculated assuming a binomial distribution for the number of positive cultures. For the datasets from Jones et al. (30) and Kohmer et al. (31), viral loads are binned into half-log_10_ intervals. Solid lines are used for models that describe the data well. **(C)** The predicted probability of transmission for a typical contact as a function of log_10_ VL given by the saturation model in Eq. [1] with *θ* = 0.20, *h* = 0.51, and *k*_m_ =8.8 × 10^2^ RNA copies (red) or by the power model in Eq. [2] with *ϕ* = 2.4 × 10^3F^ and *h* = 0.53 (blue). **(D and E)** The infectiousness profile (lines in upper panels) predicted by the infectious model assuming a saturation function (Eq.1) or a power-law function (Eq. 2), respectively. Lower panels show the relationship between the duration of the incubation period (x-axis) and estimated presymptomatic area under the infectiousness curve. Irrespective of the model used, expected presymptomatic transmission is more likely in individuals with a longer incubation period.

During a contact, the donor sheds both infectious and non-infectious viruses, and a transmission event occurs when one or more infectious viruses reach the recipient and establishes an infection (Fig. 2A). We first consider the relationship between the number of infectious viruses, *V*_*inf*_, and the measured VL, *V*, in a patient sample, e.g. a swab, using three sets of cell culture positivity data, i.e. Jaafar et al. (29), Jones et al. (30) and Kohmer et al. (31). In these three datasets, a total of 3790, 631 and 75 RT-PCR positive nasopharyngeal samples, respectively, with known cycle threshold (Ct) counts or VLs were tested for the presence of infectious virus using cell culture assay.

We examined the following three models describing the relationship between *V*_*inf*_ and *V*: 1) *V*_*inf*_ is proportional to *V*, 2) a power-law *V*_*inf*_ = _*ω*_*V*^*h*^, where *ω* and *h* are constants and 3) a Hill function: 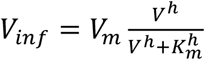, where *V*_*m*_ and *K*_*m*_ are constants. The probability of a cell culture testing positive can be expressed as *p*_*positive*_=1-exp(-*V*_*infϱ*_), where e is the probability an infectious virus will establish infection in the cell culture (see *SI Appendix*). Note that because e always appears as a product with ω or *V*_*m*_ in the expression of *p*_*positive*_, *ϱ* and the number of infectious particles, *V*_*inf*_ cannot be independently estimated from the data we used here. However, the estimated values of *h* or *k*_*m*_ describes how *V*_*inf*_ changes with *V*.

Fitting the three versions of this model to the datasets (*SI Appendix*), we found that the linear model describes all datasets poorly (Fig. 2B). The saturation model is the best model to describe the data from Jaafar et al. (Fig. 2B and Table S5), and the best fit parameter values are *h* = 0.51 and *k*_m_ =8.8 × 10^2^ RNA copies/ml (Table S6). Both the power-law model and the saturation model describe the data from Jones et al. and Kohmer et al. well (Fig. 2B). The parameter *h* is estimated to be 0.53 and 0.45, respectively (Table S6), consistent with the exponent *h* estimated from fitting the saturation model to the Jaafar et al. data. This strongly suggests that the level of infectious viruses increases sub-linearly with increases in viral load (with the exponent *h* likely being between 0.4-0.6). Because the saturation model describes all datasets well, we will mainly use this model for the analyses below. However, we reason that the evidence is not strong enough to rule out the power-law model, because the saturating behavior observed in Jaafar et al. may arise from other factors that are not part of the transmission process, such as assay noise. In addition, another study estimating transmissibility from viral load and contact tracing data did not find a saturation effect on viral load (32).

We next consider viral shedding from a donor and the establishment of infection in a recipient. We first used the saturation function above and assumed that the mean amount of infectious virus shed is proportional to the amount in a sample, *V*_*inf*_, that the exact amount is Poisson distributed and infection involves binomial sampling (see Methods). Then, the probability of one or more virions generating a successful transmission event for a typical contact at time *t* as

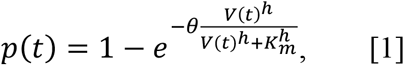

where *θ* is a composite parameter incorporating the fraction of infectious viruses reaching to the recipient and the probability of establishing an infection (*SI Appendix*). Note that when *θ* is small, *p*(*t*) can be approximated by the Hill function 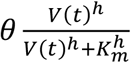. A Hill function was used previously to model the transmission probability for human immunodeficiency virus (33), influenza (34, 35), and more recently SARS-CoV-2 (21).

The values of *h* and *k*_*m*_ are determined using the cell culture data above. *θ* is a constant such that the maximum transmission probability, i.e., the maximum infectiousness, is 1 - e^−*θ*^, which is approximately θ for *θ* <<1. Multiple epidemiological studies indicate that the secondary attack rate per typical contact is low (i.e., less than 20%) (36-38). We thus set θ = 0.20 in the analysis below so that the maximum transmission probability is approximately 20% for a typical contact.

Setting *θ* = 0.20, *h* = 0.51 and *k*_*m*_ = 8.9 × 10^2^ RNA copies/ml, we calculated how infectiousness depends on viral load (Fig. 2C) and how infectiousness varies over time post-infection, *p*(*t*), i.e., the infectiousness profile, for each individual (Fig. 2D and Fig. S1). If we define the infectious period as the period when the infectiousness, *p*(*t*), is above 0.02 (i.e. 10% of the maximum probability), the infectious period ranges between 1.9 and 7.9 days with a mean of 5.5 days across thee 17 individuals (Fig. S1). For the individuals in the German dataset where the date of symptom onset is known, we calculated the presymptomatic fraction of infectiousness by dividing the area under the infectiousness curve *p*(*t*) before symptom onset by the total area under the infectiousness curve. This fraction represents the expected fraction of presymptomatic transmissions (if a person is not rapidly isolated after symptom onset). We found that the fraction ranges between 0% and 20% (Fig. 2D and Fig. S1). Interestingly, there is a statistically significant association between the duration of the incubation period, i.e., the time between infection and symptom onset, and the predicted probability of presymptomatic transmission (Fig. 2D; p-value=0.03). This suggests that the longer the incubation period, the more likely presymptomatic transmission occurs, and presymptomatic transmission is mostly driven by individuals who have an incubation period greater than 5 days.

To further cross validate this choice of parameters in the infectiousness model, we compared our model predictions with data from other epidemiological studies. First, from the infectiousness profiles predicted by our model, we calculated using Eq. [6] (see Methods) the expected serial interval for each individual (assuming random contacts) and found the mean serial interval across all 17 individuals studied to be 7.1 days. This is consistent with a mean serial interval of 6.5-8 days in the absence of active tracing and isolation efforts as estimated in Ref. (39). Second, from the infectiousness profile, we calculated using Eq. [7] (see Methods) the number of potential transmissions for each individual assuming that there are on average 13.4 typical contacts per day according to the estimates from several European countries reported in Mossong *et al*. (28). We then estimated the expected reproductive number of SARS-CoV-2 at the epidemiological level, *R*_0,*epi*_ by taking the mean of the numbers of potential transmissions. We estimated that *R*_0,*epi*_ is 5.2 for the 17 individuals (see Methods), within the range of *R*_0,*epi*_ values estimated previously for European countries (7). Therefore, these independent validations support our infectiousness model in Eq. 1.

Similarly, we derive the probability of transmission using the power-law function as

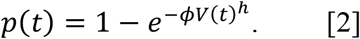

where *ϕ* is a constant. We estimated *h* = 0.53 from the data by Jones et al. (30), and *ϕ* = 0.000024 such that the model predicted mean serial interval and *R*_0,*epi*_ (6.9 days and 5.1, respectively) are consistent with epidemiological studies as was the case for the saturation model. In general, the model predictions of infectiousness are similar to the predictions using the saturation model when VL is lower than 10^7^ copies/ml; however, the predictions of the two models diverge when the VL is higher (Fig. 2C). The power function estimates similar levels of infectiousness to the estimates of the saturation model except for one individual with a high infectiousness (Fig. 2D,E, and Figs. S2 and S3). It estimates similar fraction of presymptomatic infections as the saturation model (Fig. 2E). Again, the model predicts that the fraction of expected presymptomatic transmission increases with incubation period.

Lastly, we tested whether the linear model is consistent with epidemiological data by assuming that *V*_*inf*_ is a constant fraction of *V* (Fig. S4). The model predicts that the fraction of presymptomatic infections is extremely small, i.e., less than 8% in each of the patients in the German dataset (Fig. S4B), inconsistent with epidemiological data (9, 10, 40). Therefore, datasets from cell culture experiments as well as epidemiological studies suggest that the fraction of virus particles that are infectious is not a constant over the course of infection.

### Log VL is a better surrogate measure of infectiousness than VL

There are two commonly used surrogate measures of infectiousness (11): the VL and the logarithm of VL. The total infectiousness of a person is then approximated by the area under the VL curve (AUC) and the area under the log_10_ of the VL curve (AUClog), respectively.

To identify the appropriate surrogate measure for SARS-CoV-2 infection, we first compared the predictions of these two measures with the epidemiological evidence that a large fraction (>30%) of transmissions occur during the presymptomatic stage of SARS-CoV-2 infection (9, 10, 40). Because the dates of infection and symptom onset are only available in the German dataset (23), we focused our analysis on this dataset. When AUC is used as a surrogate for infectiousness, this is very similar to using the linear model for infectiousness above. Therefore, AUC predict very small fractions of presymptomatic transmission, i.e., less than 8% in each of the patients in the German dataset, inconsistent with epidemiological data (9, 10, 40). This suggests the VL and its AUC are not good surrogates for infectiousness.

In contrast, when AUClog is used as a surrogate, we predict a sizable fraction of presymptomatic transmissions, between 2% and 27%, which is near the lower bound estimate in Ref. (10). We then correlated AUClog with the cumulative infectiousness curve calculated from the probability model based on us saturation function, i.e. Eq.1, and found that there exists a strong correlation between the two (Fig. S5A). In addition, the fractions of presymptomatic infections predicted by AUClog are very close to those predicted using the probability model (Fig. S5B). Therefore, the logarithm of VL, and its corresponding AUClog, serve as a better surrogate for infectiousness than the VL and its corresponding AUC.

### Implications for testing strategies

Using our best-fit model of how VL (Fig. 1) and infectiousness (Fig. 2D) vary with time since infection, we analyzed the impact of possible testing strategies used to reduce the potential for SARS-CoV-2 transmission. We considered two different types of tests: 1) RT-PCR, generally considered the gold standard, because of its very high sensitivity and specificity, although its performance depends on the VL and also on the quality of the sample collected (41); and 2) antigen tests, which although less sensitive, generally have faster turn-around time (minutes instead of hours to days) and can be self-administered.

We studied a hypothetical medium-sized college setting (as described in Paltiel et al. (41)). In this scenario, during a 12-week semester in a cohort of 5000 students/staff, we assume that there were 500 people infected at random times. We implemented four testing frequencies (every person every day, or every 3, 5, or 7 days), using RT-PCR or antigen testing. We assumed the sensitivity for each test varied with time since infection as in Fig. S6 (adapted from Refs. (31, 42)), and that the turnaround time was 1 day for RT-PCR and minutes for the antigen test. Given that whether infection is detected or not, as well as the time of detection, is probabilistic, for each scenario we ran 100 simulations using the best-fit model parameter values for each of the 17 individuals. We summarized the fraction of the 500 infections detected, the number of false negatives (some people may be false negatives multiple times), the average time of infection until detection, as well as the fraction of total infectiousness averted by detecting someone (assuming that person is isolated) in Fig. 3. The fraction of total infectiousness averted was defined as the area under the infectiousness curve from time of detection until resolution of infection in detected individuals divided by their total infectiousness (AUC) averaged over the 500 people infected.

**Figure 3.**
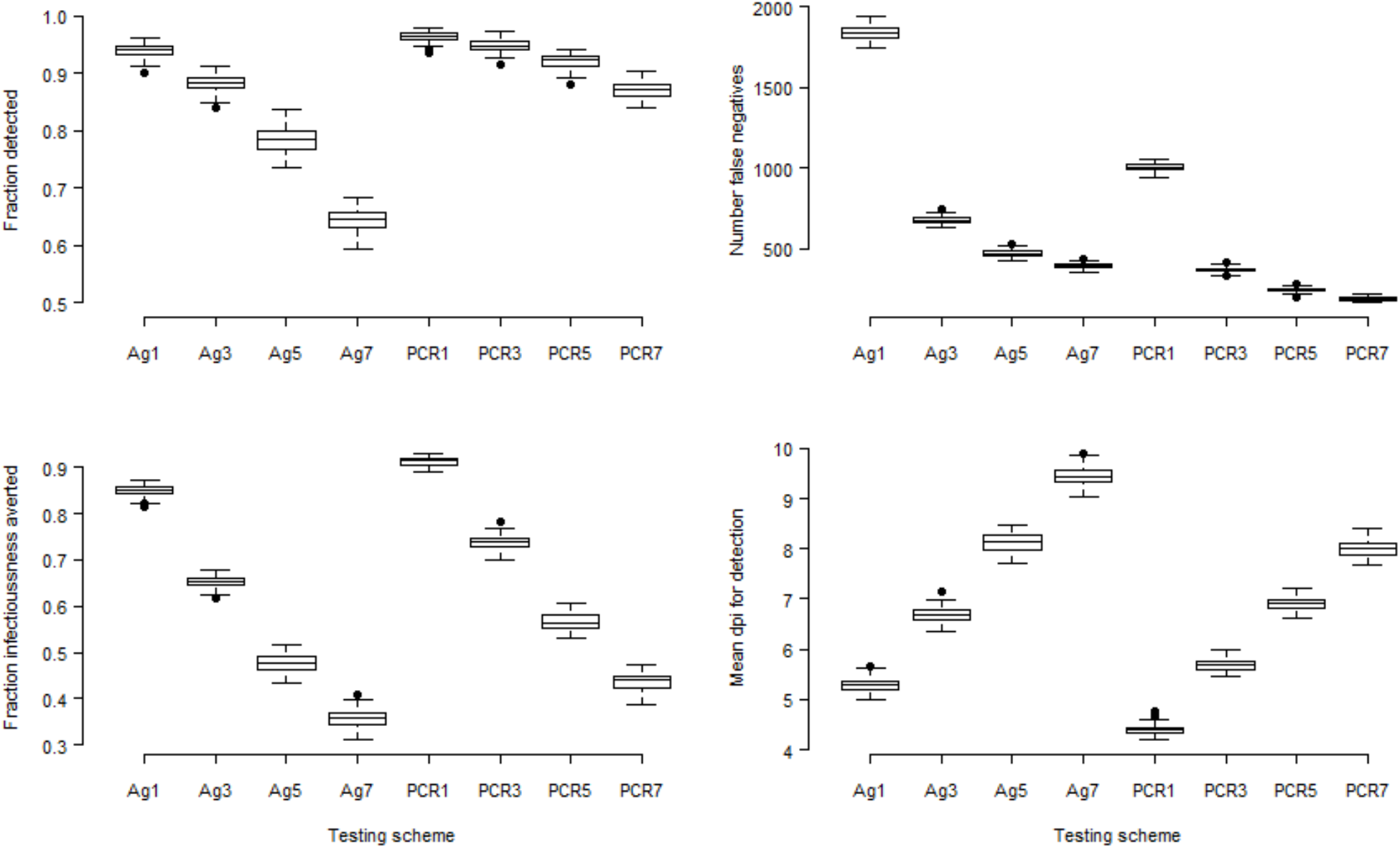
Comparison of eight testing protocols using RT-PCR or antigen tests. For each of these we considered that every person is tested every 1, 3, 5 or 7 days, as indicated in the x-axis by the number after the test type (e.g., Ag3 corresponds to antigen testing every 3 days). We plot the number of people detected (top left), the number of false negative tests (top right, note that some people may be false negatives multiple times), the fraction of total infectiousness averted (bottom left), and the average time post-infection to detection (bottom right).

We found that with a RT-PCT test, a large fraction (>80%) of infected individuals can be detected even with a testing frequency of every 7 days (Fig. 3A); whereas with an antigen test, testing at least once every 3 days is needed to achieve >80% of detection. Frequent tests (every 3 days for RT-PCR tests and every day for antigen tests) are needed to identify and isolate infected individuals early and thus avert a large fraction of infectiousness (Fig. 3C and D).

Overall, the results of these simulations show that although RT-PCR tests perform better than antigen tests in detecting infected individuals and preventing transmission, more frequent antigen testing, e.g. every day or every 3 days, is comparable to less frequent RT-PCR tests, at the expense of many more false-negative tests (Fig. 3B). This indicates that frequent antigen tests, potentially self-administered at home could be an important tool in combating spread of infection.

## Discussion

In this study, we constructed mathematical models to describe the VL kinetics of SARS-CoV-2 in the upper respiratory tract, and their relationship with the infectiousness of an individual. Fitting a viral dynamic model that included an innate immune response to data from Refs. (23) and (25), we estimated several key parameter values. The death rate of productively infected cells was estimated to be around 1.7 d^-1^. Thus, once infected cells start producing viruses they live on average 0.6 days. We estimated the mean within-host reproductive number, R_0_,within, in the URT to be 7.4 with variation among individuals examined, ranging between 2.6 and 14.9. For individuals with known dates of infection and symptom onset, we found that longer incubation periods had higher potential for presymptomatic transmission. A similar finding was reported in a recent study estimating the fraction of presymptomatic transmissions by the duration of the incubation period from transmission pair data (43).

To model viral transmission, we estimated the relationship between the number of infectious viruses in a sample and the sample VL by fitting models to three datasets on infectious virus cell culture positivity (29-31). This led to several interesting findings. First, a consistent finding across the three datasets was that the number of infectious viruses does not increase linearly with increases in viral load, suggesting viral load itself or the area under the viral load curve is not a good surrogate for infectiousness. Instead, we found that the number of infectious viruses increases sub-linearly with increases in viral load. This makes log VL or area under the log VL curve good surrogates for infectiousness. It is not clear what causes this sublinear relationship. One possible mechanism is the development of neutralizing antibodies when VL is high (44), which renders some viruses non-infectious. Further experiments are needed to understand this sublinear relationship. Second, a saturation effect on the infectious viruses when viral load is very high, e.g., >10^9^ copies/ml, is needed to explain data from Jaafar et al. (29); however, saturation is not needed to explain the data from Jones et al. (30) and Kohmer et al. (31). The saturation effect, if present, could be due to assay inaccuracies at very high VLs or could arise from processes in vivo such as viral neutralization due to neutralizing antibodies in high VL samples. In fact, van Kampen et al. (45) showed that a neutralizing antibody titer of 1:20 or more in hospitalized patients was independently associated with the isolation of non-infectious SARS-CoV-2 in the respiratory tract. This inconsistency in results vis à vis saturation leads to uncertainties in predicting infectiousness when viral load is very high. Further experiments measuring the infectious virus concentration especially from samples with high VLs is needed to address this issue. In our study, irrespective of the model used, we found that the risk of transmission for a typical contact of relative short duration becomes high when the VL exceeds between 10^6^ and 10^7^ RNA copies/ml. This is consistent with the results from Wolfel et al. (23), where infectious viruses were recovered only when VL exceeded 2×10^5^ RNA copies/swab and the results from Ref. (46) where infectious virus was mainly isolated from specimens with ≥ 10^6^ virus N gene copies/ml. The results are also consistent with the findings in van Kampen et al. (45) where in hospitalized patients with COVID-19 VLs > 10^7^ copies/ml were associated with isolation of infectious virus.

Using the predicted infectiousness over time for each individual, we evaluated the effectiveness of two testing platforms: RT-PCR and antigen tests. RT-PCR tests are highly sensitive; however, they are costly and may take days to obtain the result. On the other hand, antigen tests are less sensitive, but are easy to administer and provide results in less than an hour. Our modeling suggests that RT-PCR tests are better than antigen tests at both detecting infected individuals and effectively reducing total infectiousness when testing is used as a tool for safe reopening of schools and workplaces. However, when frequent RT-PCR testing, say every 7 days, is not feasible due to its high cost and complexity in properly administering these tests, more frequent antigen tests (i.e., every 1-3 days) could be used instead; however, this will lead to higher number of false negative results due to the large number of antigen tests performed.

There are limitations to our models. First, the data we used for model inference were from infected individuals with relatively mild or no symptoms (23, 25), who rapidly cleared the virus. The parameter values and relationships we estimated between VL and infectiousness thus may be biased towards mildly symptomatic and asymptomatic individuals. Further work is needed to extend our analysis to individuals with different levels of symptom severity (47). However, we note that people with severe symptoms will likely often be quarantined and contribute less to the spread of the virus. Second, the relationship between VL and the number of infectious particles is inferred from data aggregated from many individuals, and thus it assumes homogeneity across individuals. Further work measuring individual level heterogeneity in the relationship between infectious viral shedding and VL will help to characterize heterogeneity in individual infectiousness and help make more precise predictions of the impact of testing strategies on transmission.

Overall, our model linking within-host VL dynamics to infectiousness provides a crucial tool for evaluating both non-pharmaceutical and pharmaceutical interventions, and aiding public health policy decisions (48). For example, there is an emerging need to quantify the extent of transmission of asymptomatic individuals and particularly of school-aged children (49), who may have a similar range of VLs as adults (30). Administration of vaccines or effective therapeutics may lead to reduced VLs in the upper respiratory tract (16). Our model will help to quantify the impact of vaccination on the infectiousness of a person.

## Methods

### Target cell limited (TCL) model

We first study a within-host model based on target cell limitation (TCL). The model keeps track of the total numbers of target cells (*T*), cells in the eclipse phase of infection (*E*), i.e., infected cells not yet producing virus, productively infected cells (*I*) and viruses measured in swab samples (*V*). The ordinary differential equations (ODEs) describing the model are

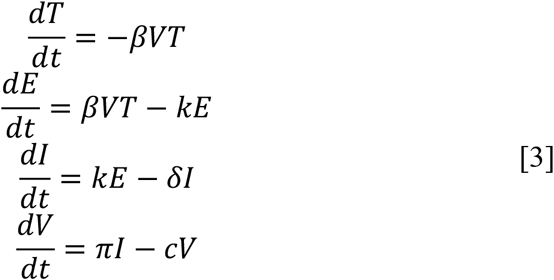

In this model, target cells are infected by virus with rate constant *β*. Cells leave the eclipse phase and become productively infected at per capita rate *K*. Productively infected cells die at per capita rate *δ*. We use *V* to describe viruses measured in pharyngeal swabs, which are a proportion of the total virus in the URT. Therefore, the rate, *π*, is the product of the viral production rate per infected cell and the proportion of virus that is sampled in a swab. Viruses are cleared at per capita rate *c*. See *SI Appendix* for further details.

From this model, we calculate the within-host reproductive number for SARS-CoV-2, *R*_0,*within*_ as

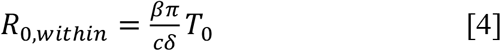

where *T*_0_ is the initial number of target cells.

### Innate response model

We extend the TCL model by including a prototypical innate response (e.g., type-I interferon) following the framework presented in previous models for influenza infection dynamics (26, 27, 50). Immune mediators are produced from infected cells and bind to receptors on target cells stimulating an antiviral response that makes cells refractory to viral infection (*R*). Such cells are said to be refractory cells or cells in an antiviral state (51). In addition to the compartments in the TCL model, the innate response model keeps track of cells refractory to infection (*R*). For simplicity and due to a lack of data, we do not explicitly consider the specific immune mediators (e.g., cytokines) or their concentration. Instead, we make the quasi-steady-state assumption that the dynamics of these mediators are fast and thus their concentration is proportional to the number of infected cells (see *SI Appendix* for details).

The ODEs for the innate response model are

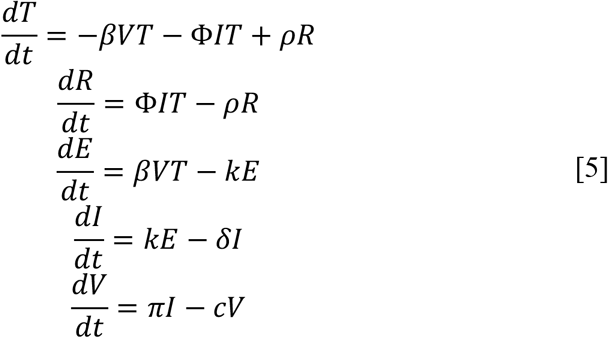

where Φ is a constant describing the rate that innate signaling makes a target cell refractory and *ρ* is the rate that refractory cells transition back into target cells.

### Data, estimating time of infection, parameter fitting and analysis

For the German dataset, we digitalized longitudinal VL data from throat swabs of the 9 infected individuals reported in Wolfel et al. (23). The infected individuals are young to middle-aged professionals, without underlying disease, who were identified because of known close contact with an index case. All patients were hospitalized but had a comparatively mild clinical course of disease. For the NBA dataset, we used data reported in Kissler et al. (25). We included 9 individuals for whom multiple detectible VL measurements were available before the viral peak. Note that VLs were reported in copies/swab by Wolfel et al. (23) and in copies/ml in Kissler et al. (25). Since we did not find significant difference in parameter estimates between the two datasets (see main text), the unit of choice/reporting may not strongly impact our results. For consistency, we use copies/ml as the reporting unit.

We use a population approach, based on non-linear mixed effect modeling (unless specified otherwise), to fit the model simultaneously to VL data from the two datasets, using the software Monolix (Lixoft, SA, France). We calculated correlations between the incubation periods and the fractions of predicted presymptomatic transmission using Pearson correlation.

### The model for infectiousness

To calculate the probability of transmission given a typical contact of duration *𝜏*, we assume that 𝜏 is small enough (on the order of minutes or hours) that the total VL in the URT of the donor and thus the level of infectious viruses, *V*_*inf*_ is approximately constant during the contact between time *t* and *t* + *𝜏*. We then assume that the number of infectious viruses shed per unit time is *μV*_inf_, where μ is a constant. Of these, a fraction, *φ*, reaches the URT of the recipient. Then on average the total number of infectious viruses reaching the recipient for a contact of duration *τ*, is n = *φμτV*_*inf*_ Airborne pathogens tend to be randomly distributed in the air (52). Thus, we assume the number of infectious viruses reaching the recipient during a contact is a random variable *X* that is Poisson distributed with parameter *n*. We further assume that each infectious virus that reaches the recipient has a probability *v* to successfully establish infection and that if *X* viruses reach the recipient the probability to establish an infection is given by the binomial distribution Bin(*X, v*). However, since *X* follows a Poisson distribution one can show the distribution of the number of viruses that successfully establish an infection follows a Poisson distribution with parameter 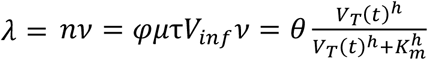, for the saturation model where *θ* = *φμτV*_*m*_*v*. Then, the probability of one or more virions generating a successful transmission event for a typical contact at time *t* is given by Eq. [1].

### Estimating the expected serial intervals and *R*_0,*epi*_ from infectiousness profiles

To calculate the expected serial interval (or the generation interval), we assume that contacts are randomly distributed over time. Then the expected serial interval for the *i*^th^ individual, *SI*_*i*_, can be calculated as

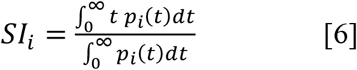

where *p*_i_(*t*) is the probability of transmission (Eq. 1) given a typical contact for individual *i*. The mean serial interval across all individuals in our study is calculated as the mean of the *SI*s calculated for individuals in the two datasets.

To calculate the expected epidemiological reproductive number, we assume that there are on average 13.4 contacts of a relatively short duration, per day according to the estimates in Mossong *et al*. (28). Then the expected epidemiological reproductive number for individual *i* is calculated as

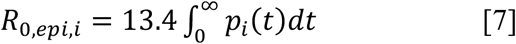

The mean epidemiological reproductive number across all individuals in the two datasets, *R*_0,*epi*_ is calculated by taking the average of *R*_0,*epi,i*_ across all individuals.

Note that the calculation of *R*_0,*epi*_ above is a rough approximation, because it implicitly makes the simplifying assumption that contacts are randomly distributed over time and every individual has the same number of contacts per day. This is used in our study to show that the choice of parameter values (for *θ, h* and k_*m*_) are broadly consistent with estimates of epidemiological parameters such as the mean serial interval and *R*_0,*epi*_ However, it should not be treated as an exact expression. See Ref. (53) for discussion of formally calculating *R*_0,*epi*_ in the context of SARS-CoV-2 transmission.

### Model and assumptions for evaluating testing strategies

Several studies have remarked that testing sensitivity in clinical practice can be much lower than the theoretical detection limit would indicate. For example, Kucirka et al. (42) suggested that the sensitivity of a RT-PCR test depends on the time since infection (a reflection of the VL) and that it is never more than 80%. Although there are many RT-PCR test platforms and protocols in use, the general sensitivity over infection stages is likely not substantially different. To examine testing protocols under the best of circumstances, we assume much better performance for RT-PCR tests than suggested by Kucirka et al. (42), with no detection if the VL is below 10^3^ copies/ml, but 90% sensitivity for any VL above that (Figure S4A). We compare this test, with an antigen test with characteristics as presented in Kohmer et al. (31), who compared the performance of several antigen tests with the results of RT-PCR. Based on their data for the SARS-CoV-2 Rapid Antigen Test (Roche Diagnostics) versus the VL in the sample, we fit the performance of the test to a logistic type relation between VL and positivity detection yielding the curve shown in Figure S5B (see the *SI Appendix* for further details). An infected person’s probability of being detected is a Bernoulli trial based on the sensitivity of the test (as in Fig. S4).

## Data Availability

All data are available in the manuscript

## Acknowledgements

Portions of this work were done under the auspices of the U.S. Department of Energy through Los Alamos National Laboratory, which is operated by Triad National Security, LLC for the National Nuclear Security Administration of the U.S. Department of Energy (contract No. 89233218CNA000001). The work was supported by the Laboratory Directed Research and Development program of Los Alamos National Laboratory (projects No. 20200743ER, 20200695ER, and 20210730ER), by NIH grants R01-AI028433, R01-OD011095 (ASP), R01-AI15270301 (RK) and R01-AI116868 (RMR), by the National Science Foundation RAPID grant PHY-2031756 (ASP), by the Defense Advanced Research Projects Agency (contract No. HR0011938513) and by the DOE Office of Science through the National Virtual Biotechnology Laboratory, a consortium of DOE National Laboratories focused on response to COVID-19, with funding provided by the Coronavirus CARES Act. This work was competed at the Aspen Center for Physics, which is supported by National Science Foundation grant PHY-1607611.

