## Supplementary Material for "In vivo kinetics of SARS-CoV-2 infection and its relationship with a person’s infectiousness"

SI Appendix for

#### Affiliations:

**This PDF file includes:**

Supplementary Methods
SI references
Figures S1 to S6
Tables S1 to S6

#### Supplementary Methods

##### 1. Choice of fixed parameter values

Total target cell numbers in the absence of infection ($T_0$)

It has been estimated that there are $4\times10^8$ epithelial cells in the upper respiratory tract (URT) (1). Hou et al. recently estimated that the fraction of cells that express angiotensin-converting enzyme 2 (ACE2), i.e. the receptor for SARS-CoV-2 entry, on cell surface is approximately 20% in the URT (2). There is a much higher fraction of cells expressing the type II transmembrane serine protease TMPRSS2, a co-receptor for SARS-CoV-2 entry (2). Therefore, in our model, we assume the initial number of target cells in the URT, $T_{1,0}$ $=8\times10^7$ cells, i.e., 20% of the total epithelial cells. Note that for a standard viral dynamics model (as well as in our immunity model described below), the number of initial target cells and the virus production rate are unidentifiable and only their product is identifiable (3). Thus, an increase (decrease) in the initial number of target cells will lead to corresponding decrease (increase) in the estimate of the virus production rate but not in the estimate of other parameters such as $\delta$ or $R_0$ (see Eqs. [4] and [5]).

Initial number of infected cells (in an eclipse phase), $E_0$

Evidence strongly suggests that the URT is the initial site of infection (2, 4). Thus, we assume that one cell in the URT is infected at the start of infection, $I_0 = 1$ cell, rather than setting an initial viral load, which avoids the complication of predicting whether one or more infectious virions would be present for any chosen $V_0$. This approach is similar to that in Ref. (5), which showed that this assumption does not change the dynamics of the model significantly as any initial viral particles that succeed in initiating infection must infect one or more cells (rapidly) before being cleared. In a sensitivity analysis, we test $I_0 = 10$ cells.

Virus clearance rate, c

We set c=10/day, because *in vivo* viral clearance is usually fast in many infections, including for respiratory infections such as influenza (1, 5, 6). We and others have used this value of $c$ in previous models of infection by SARS-CoV-2 (7, 8). In sensitivity analyses, we set c= 5 or 20/day.

Eclipse period, $1/k$

We set $k = 4$ /day (corresponding to $\frac{1}{k}$ =6 hours) according to cell culture experiments suggesting infected cells start to produce virus between 4-8 hours post infection (9). In sensitivity analyses, we set $\frac{1}{k}$ =4 or 8 hours, with the 4 hour choice motivated by the experiments in Hou et al. (2) that showed viral titers of $10^4$ PFU/ml in vitro at 6 hour post infection, the earliest time point sampled.

##### 2. The target cell limited (TCL) model

We first construct a within-host model based on the target cell limited model. The model keeps track of the total numbers of target cells ($T$), cells in the eclipse phase of infection ($E$), i.e., infected cells not yet producing virus, productively infected cells ($I$) and total viruses ($V_{URT}$). To compare the model with data, we keep track of sampled viruses, i.e., virus levels measured in nasal pharyngeal swabs, $V$, and assume that these levels are proportional to the actual number of viruses in the URT, $V_{URT}$. The ordinary differential equations (ODEs) describing the model are

$$
\begin{aligned}
\frac{dT}{dt} &= -\beta_{URT}V_{URT}T \\
\frac{dE}{dt} &= \beta_{URT}V_{URT}T - kE \\
\frac{dI}{dt} &= kE - \delta I \\
\frac{dV_{URT}}{dt} &= pI - cV_{URT}
\end{aligned}
\qquad [S1]
$$

$$V = fV_{URT}$$

where $f$ is the proportion of virus sampled from the URT in either a single swab for the German data or in 1 mL of fluid that the swab was placed in for the NBA data.

Model simplification.
From Eqn. S1, we have:

$$\frac{dV}{dt} = f\frac{dV_{URT}}{dt} = fpI - cV$$

Let $\pi = fp$, and $\beta = \beta_{URT}/f$, we get we get the following ODEs shown as Eqn. [2] in the main text:

$$
\begin{aligned}
\frac{dT}{dt} &= -\beta VT \\
\frac{dE}{dt} &= \beta VT - kE \\
\frac{dI}{dt} &= kE - \delta I \\
\frac{dV}{dt} &= \pi I - cV
\end{aligned}
\qquad [S2]
$$

#### 3. Innate immunity models

We constructed three versions of the innate immune model. The first version of the model is used in the analyses in the main text, and is termed the innate immune model throughout the main text.

##### a. *Innate immune model - refractory cells*

In the first version of the innate immune model, we keep track of type I interferon (*F*) and cells refractory to infection (*R*), in addition to the compartments in the TCL model. We assume that binding of interferons to receptors on target cells stimulates genes that make target cells refractory to infection

The full ODEs for target cells, refractory cells and interferon are

$$
\begin{aligned}
\frac{dT}{dt} &= -\beta VT - \phi FT + \rho R \\
\frac{dR}{dt} &= \phi FT - \rho R
\end{aligned}
\qquad [S3]
$$

$$\frac{dE}{dt} = \beta VT - kE$$
$$\frac{dI}{dt} = kE - \delta I$$
$$\frac{dV}{dt} = \pi I - cV$$
$$\frac{dF}{dt} = sI - \mu F$$

In this model, the impact of the innate immune response is to convert target cells into refractory cells at rate $\phi FT,$ where $\phi$ is a rate constant. Refractory cells can become target cells again at rate $\rho$. Interferon is produced and cleared at rates $s$ and $\mu$, respectively.

To minimize the number of unknown parameters, we simplify the model by making the quasi-steady-state assumption that the interferon dynamics are much faster than the dynamics of infected cells and assume that $\frac{dF}{dt} = 0$. Thus $sI = \mu F$ or $F = \frac{s}{\mu} I$.

Let $\Phi = \phi \frac{s}{\mu}$, so that the ODEs for the innate immunity model become

$$\frac{dT}{dt} = -\beta VT - \Phi IT + \rho R$$
$$\frac{dR}{dt} = \Phi IT - \rho R$$
$$\frac{dE}{dt} = \beta VT - kE \qquad \text{[S4]}$$
$$\frac{dI}{dt} = kE - \delta I$$
$$\frac{dV}{dt} = \pi I - cV$$

**b.** *Innate immune model – reducing infectivity*

The second version of the model considers that interferons may reduce infectivity, i.e., make cells less susceptible to infection. Again, we make the quasi-steady-state assumption that the interferon dynamics are much faster than the dynamics of infected cells and assume that $F$ is proportional to $I$. The ODEs for the model are

$$\frac{dT}{dt} = -\frac{\beta}{1 + \gamma I} VT$$
$$\frac{dE}{dt} = \frac{\beta}{1 + \gamma I} VT - kE$$
$$\frac{dI}{dt} = kE - \delta I \qquad \text{[S5]}$$
$$\frac{dV}{dt} = \pi I - cV$$

where $\gamma$ is a constant representing the effect of innate response mediators such as type I interferon.

**c.** *Innate immune model – reducing virus production*

The third version of the model considers the potential impact of the innate response on reducing virus production from infected cells. For example, in the hepatitis C virus infection administration of type I interferon reduces viral RNA replication and viral production (10, 11). As above, we make the quasi-steady-state assumption that the interferon dynamics are much faster than the dynamics of infected cells and assume that $F$ is proportional to $I$. The ODEs for the model are

$$\begin{aligned} \frac{dT}{dt} &= -\beta VT \\ \frac{dE}{dt} &= \beta VT - kE \\ \frac{dI}{dt} &= kE - \delta I \\ \frac{dV}{dt} &= \frac{\pi}{1+\gamma I} I - cV \end{aligned} \qquad \text{[S6]}$$

where $\gamma$ is a constant representing the effect of innate response mediators such as interferon.

#### 4. Model fitting and parameter estimation

Estimating time of infection

To estimate the times of infection of individuals in the NBA dataset, we fit both the TCL model and the innate immune model to viral load measurements from each individual by minimizing the least-squared residual error between viral load measurements and the model predicted viral load on a logarithm scale. The best-estimates of the times of infection are reported in Table S1.

Parameter estimation from all datasets

We used a population approach, based on non-linear mixed effect modeling, to fit the data from all patients simultaneously, with each of the models. We fixed infection dates as estimated using that model (Table S1) for the NBA dataset and to the known infection dates of the German dataset. We allowed random effects on the fitted parameters.

We analyzed the source of the dataset, i.e., the NBA data or the German data, as a categorical covariate for fitted parameters. We first tested the model by assuming all of the parameters covary with the covariate. We then exclude the parameter that has the lowest p-value by the Pearson's correlation test, which tests whether covariates should be removed from the model by Monolix. All estimations were performed using Monolix (Monolix Suite 2019R1, Antony, France: Lixoft SAS, 2019. lixoft.com/products/monolix/).

#### 5. Inferring the relationship between the number of infectious viruses and viral load

To understand how the level of infectious viruses relates to viral load, we constructed and fit mathematical models to the three datasets. In the first dataset, 'the Jaafar dataset', Jaafar et al. measured the cycle threshold (Ct) values using RT-PCR and infectious virus positivity using cell culture assay from a total of 3790 RT-PCR positive samples (12). In the second dataset, 'the Kohmer dataset', Kohmer et al. reported Ct values and their corresponding viral loads and infectious virus positivity for a total of 75 RT-PCR positive samples (13). In the third dataset, 'the Jones dataset', Jones et al. analyzed 631 RT-PCR positive samples by cell culture. Below, we first show the calculation of viral loads from the Ct values reported in Jaafar et al., and then show the derivation and model fits to the three datasets.

*Relationship between viral load and Ct values reported in Jaafar et al.* (12)
First, the viral load, $V$, measured as RNA copies in a sample is related to cycle threshold values, $C$, as:

$$V = a\exp(-bC) \qquad \text{[S7]}$$

where the constants $a$ and $b$ are determined by the RT-PCR assay used. Jaafar et al. measured the Ct values using the LightCycler 480 instrument (Roche Diagnostics) (12). According to a recent report (14), $a = 1.441 \times 10^{14}$ and $b = 0.685$, for this instrument. Thus,

$$V = 1.441 \times 10^{14} e^{-0.685C} \qquad \text{[S8]}$$

Note that the exponential term, $e^{-0.685C}$, can be written as $1.98^{-C}$, where the base of this power term is 1.98, i.e., very close to 2, the factor one would expect viral RNA to increase by during each PCR cycle if the PCR amplification is perfect.

*Relationship between viral load and infectious viruses*
We assume the number of infectious viruses that was in the sample for cell culture experiment to be a random variable, $Y$, that follows a Poisson distribution. We consider three alternative models describing how the mean number of infectious viruses in a sample, $V_{inf} = E(Y)$, is related to viral load measured by qPCR: the 'linear' model, the 'power-law' model and the 'saturation' model:

1. The linear model

We first assume that the mean of the infectious virus in a sample, $V_{inf}$, is proportional to the viral load, $V$, in the sample, i.e.,

$$V_{inf} = E(Y) = AV \qquad \text{[S9]}$$

This is the simplest model describing the relationship between infectious viruses and viral load. However, as we will show below, the model does not fit the three datasets. Therefore, we developed two additional models to describe this relationship.

2. The power-law model

In this model, we assume that the mean of the infectious virus in a sample, $V_{inf}$, is related to the viral load, $V$, by a power-law function

$$V_{inf} = E(Y) = BV^h \qquad \text{[S10]}$$

where $B$ and $h$ are constants.

3. The saturation model

In this model, we assume that the mean of the infectious virus in a sample, $V_{inf}$, is related to the viral load, $V$, by a Hill function

$$V_{inf} = E(Y) = V_m \frac{V^h}{V^h + K_m^h} \qquad \text{[S11]}$$

where $V_m$ and $K_m$ are constants.

*Probability of cell culture positivity*

We now calculate the probability of cell culture positivity for the three models in Eqs. [S9]-[S11]. We assume the number of infectious viruses in the sample, *Y*, is a random variable that follows a Poisson distribution with mean *E[Y]*. If each infectious virus has a probability $\varrho$ to establish infection such that the cell culture becomes positive, then the number of viruses that successfully establish an infection in cell culture is Poisson with parameter $\lambda = E(Y)\varrho = V_{inf}\varrho$. Thus, the probability of one or more viruses successfully infecting the culture so that it tests positive is

$$p_{positive} = 1 - \exp(-\lambda) = 1 - \exp(-V_{inf}\varrho) \qquad \text{[S12]}$$

Using the subscript $i$ to denotes the model for $V_{inf}$, for the linear model, we substitute the expression for $V_{inf}$ in Eq. [S9] into Eq. [S12], and get

$$p_{positive,1}(V) = 1 - \exp(-AV) \qquad \text{[S13]}$$

where $D = A\varrho$.

For the power-law model, we substitute the expression in Eq. [S10] into Eq. [S12], and get

$$p_{positive,2}(V) = 1 - \exp(-GV^h) \qquad \text{[S14]}$$

where $G = B\varrho$.

For the saturation model, we substitute Eq. [S11] into Eq. [S12], and get

$$p_{positive,3}(V) = 1 - \exp\left(-J\frac{V^h}{V^h + K_m^h}\right) \qquad \text{[S15]}$$

where $J = V_{max}\varrho$.

*Model fitting*

Jaafar et al. reported the total number of samples and the number of samples that were positive in cell culture for each Ct value (ranging between 11 and 37) (12). We can calculate the likelihood of observing these numbers given the probabilities of cell culture positivity as defined in Eqs. [S13]-[S15]. Then the probability of observing the $m_j$ positive cell cultures in a total of $n_j$ cultures where $j$ denotes the $j^{th}$ Ct value of the sample put into culture, $j$= 11, 12…,37, is

$$p_{i,j} = \binom{n_j}{m_j} p_{positive,i}(V_j)^{m_j}\left(1 - p_{positive,i}(V_j)\right)^{n_j - m_j} \qquad \text{[S16]}$$

where $V_j$ is the viral load corresponding to the $j^{th}$ Ct value.

The negative log-likelihood (NLL) of the $i^{th}$ model given all the data in Jaafar et al. is then given by

$$NLL_i = -\sum_j \log p_{i,j}\,, \quad i = 1,2,3 \qquad \text{[S17]}$$

In the Kohmer dataset and the Jones dataset, for each sample, the viral load and the cell culture positivity were reported. We calculate the likelihood of the k[th] observation being positive or negative as

$$p_{i,k} = \begin{cases} p_{positive,i}(V_k), & if\ the\ kth\ observation\ is\ positive \\ 1 - p_{positive,i}(V_k), & if\ the\ kth\ observation\ is\ negative \end{cases} \qquad \text{[S18]}$$

where $V_k$ is the viral load of the $k^{\text{th}}$ observation.

The negative log-likelihood (NLL) of the $i^{\text{th}}$ model given the Kohmer dataset is then given by

$$NLL_i = -\sum_k \log p_{i,k}\,, \quad i = 1,2,3 \qquad \text{[S19]}$$

*<u>Model comparison</u>*

To compare models, we compute the AIC scores as

$$AIC_i = 2k_i + 2NLL_i, \quad i = 1,2,3 \qquad \text{[S20]}$$

where $k_i$ is the number of estimated parameters of the $i^{\text{th}}$ model, i.e., $k_1 = 1$ (parameter *A*), $k_2 = 2$ (parameters *D* and *h*) and $k_3 = 3$ (parameters *G*, *h* and $K_m$).

*<u>Results</u>*

We fitted the three models to the three datasets by minimizing the NLLs described in Eqs. [S17] and [S19]. According to the AIC scores, with the lower the score the better the model, the saturation model is the best model to describe the Jaafar dataset (Table S5). The power-law model is the best model to describe both the Jones dataset and the Kohmer dataset (Table S5). The saturation model only had slightly higher AIC scores and thus also has considerable support (15).

Interestingly, the estimated parameter values using the saturation model are very similar across the two datasets (Table S6), emphasizing the reliability of these estimates. For the saturation model iin the main text, we use $h = 0.51$, $K_m = 8.8 \times 10^6$ RNA copies/ml as estimated from the Jaafar dataset. For the power-law model in the main text, we use $h = 0.53$ as estimated from the Jones dataset.

### Supplementary Figures

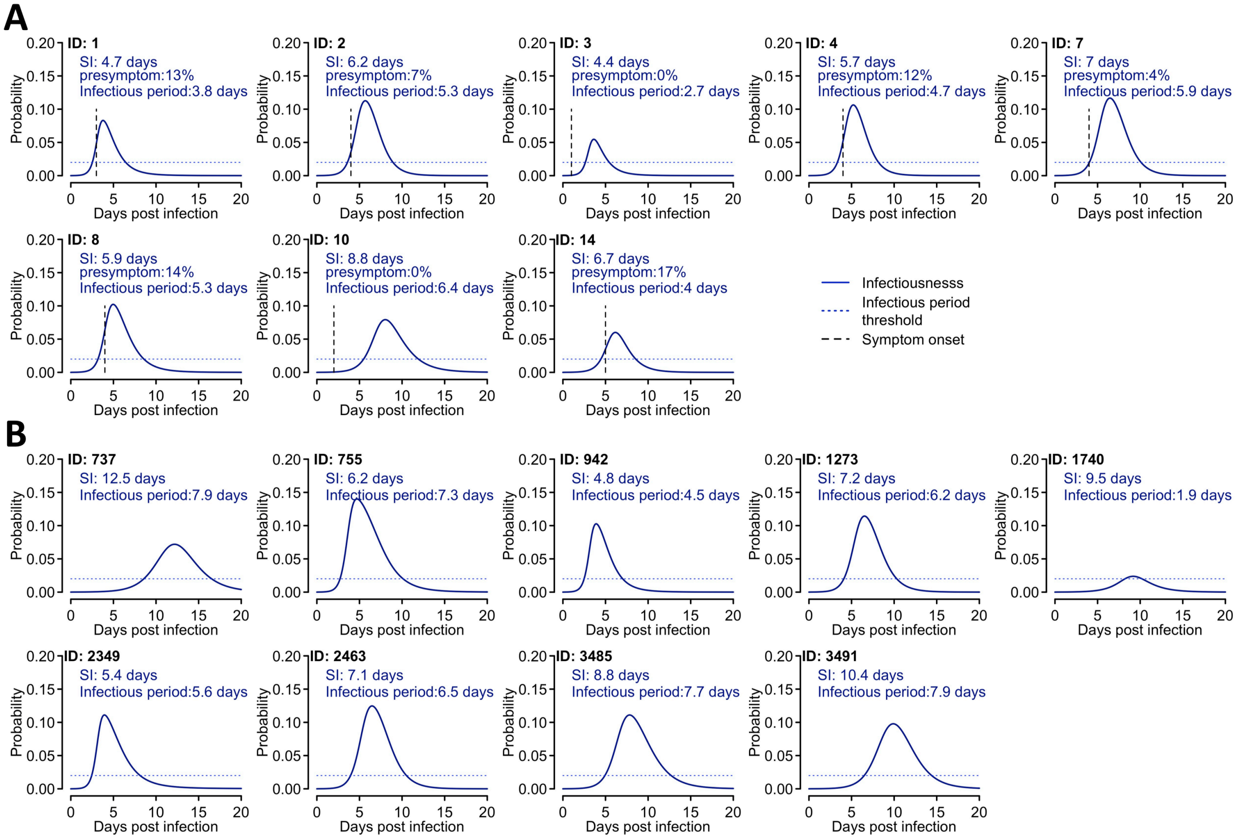

**Figure S1. The individual infectiousness profile (blue lines) predicted by the saturation model for individuals in the Germany study (A) and the NBA study (B)**. Parameters used are the same as in Fig. 2D. In panel A, the expected serial interval (SI), the fraction of presymptomatic transmission and the infectious period are reported. In panel B, only the expected serial interval (SI) and the infectious period are reported., because the symptom onset dates for these individuals are unknown. Horizontal dashed lines denote the threshold we defined (i.e. 0.02) above which a person becomes infectious. Vertical lines in panel A denote the time of symptom onset as reported in Ref. (16).

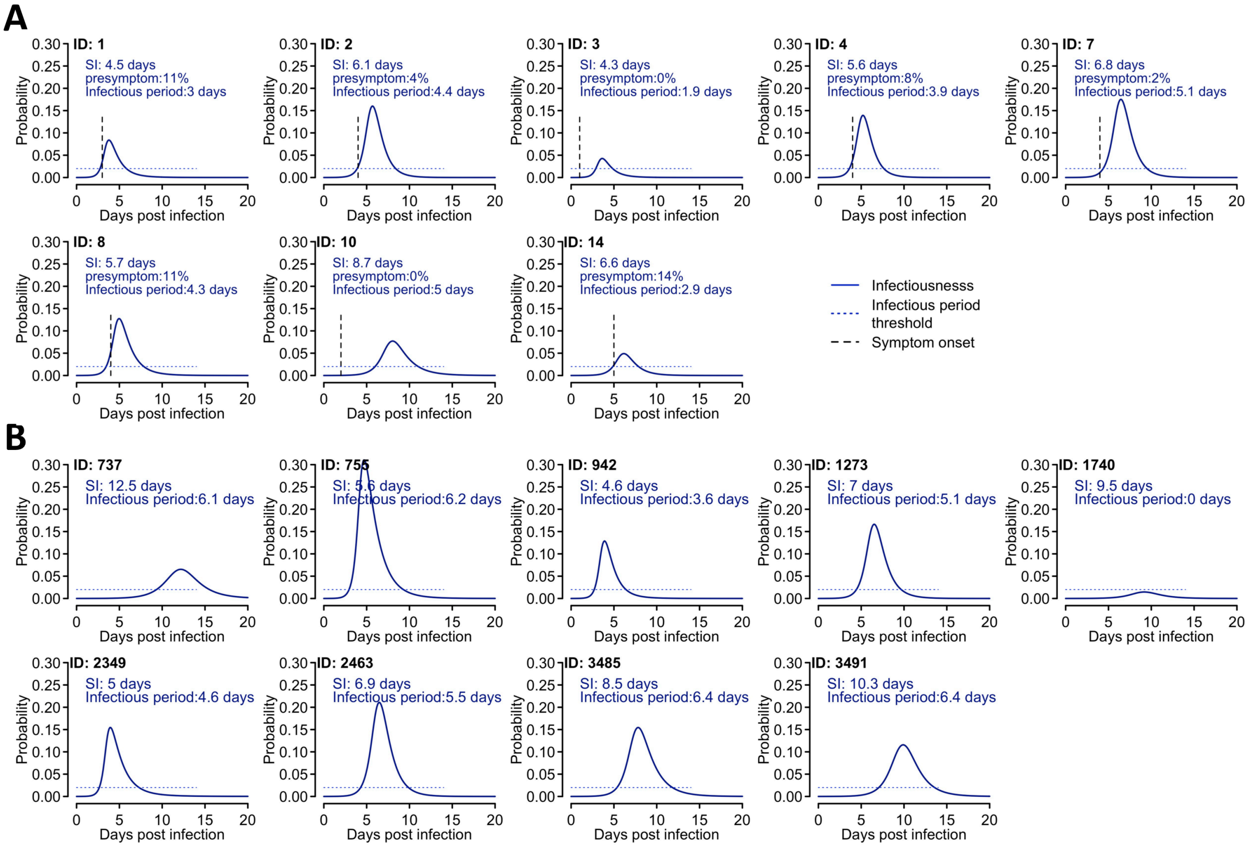

**Figure S2. The individual infectiousness profile (blue lines) predicted by the power-law model for individuals in the Germany study (A) and the NBA study (B)**. Parameters used are the same as in Fig. 2E. Other notations are the same as Fig. S1.

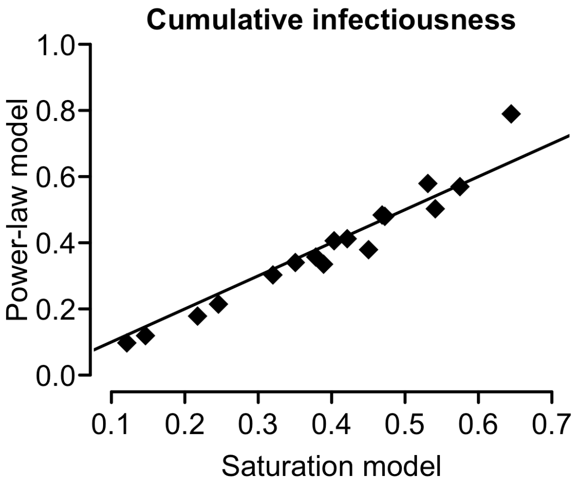

**Figure S3. Predictions of the cumulative infectiousness of individuals (dots) using the saturation model (Eqn. 1 of the main text) and the power-law model (Eqn. 2 of the main text).** The black line denotes the line of y=x.

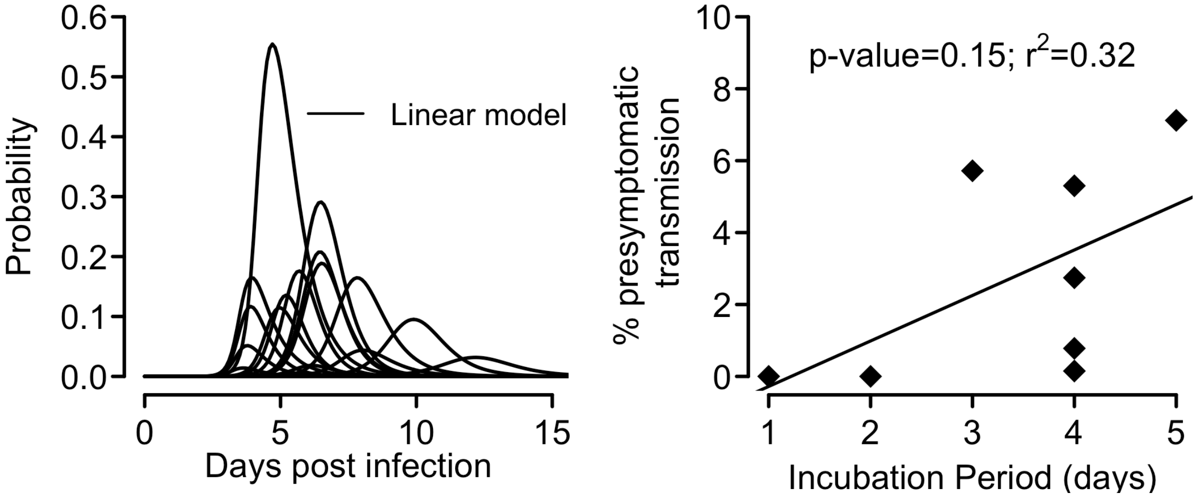

**Figure S4. The infectiousness profile (A) predicted by the infectious model assuming a linear function, and the relationship between the duration of the incubation period (x-axis) and the presymptomatic area under the infectiousness curve estimated assuming a linear function (B).**

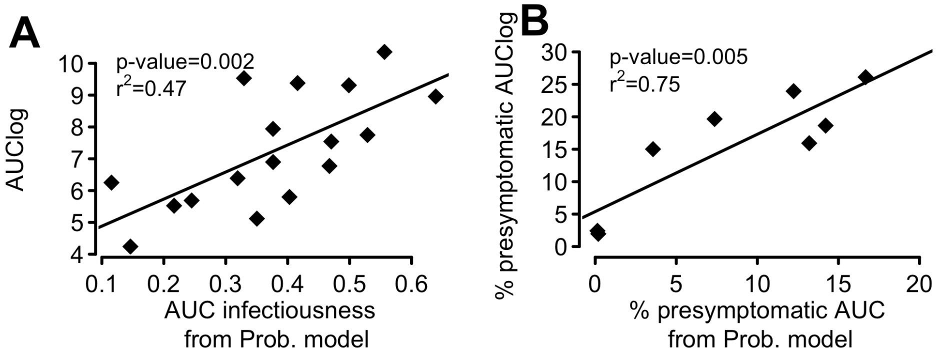

**Figure S5. Consistency between the area under the logarithm of the viral load curve, i.e. AUClog, and the area under the infectiousness curve (AUC infectiousness). (A)** Regression of and correlations between the area under the curve of infectiousness from the probability model, $p(t)$, and AUClog for all the 17 individuals studied. **(B)** Regression of and correlations between the percentage of presymptomatic transmission predicted by AUClog calculated from the model fit and by the AUC of the infectiousness curve for the 8 individuals in the German study.

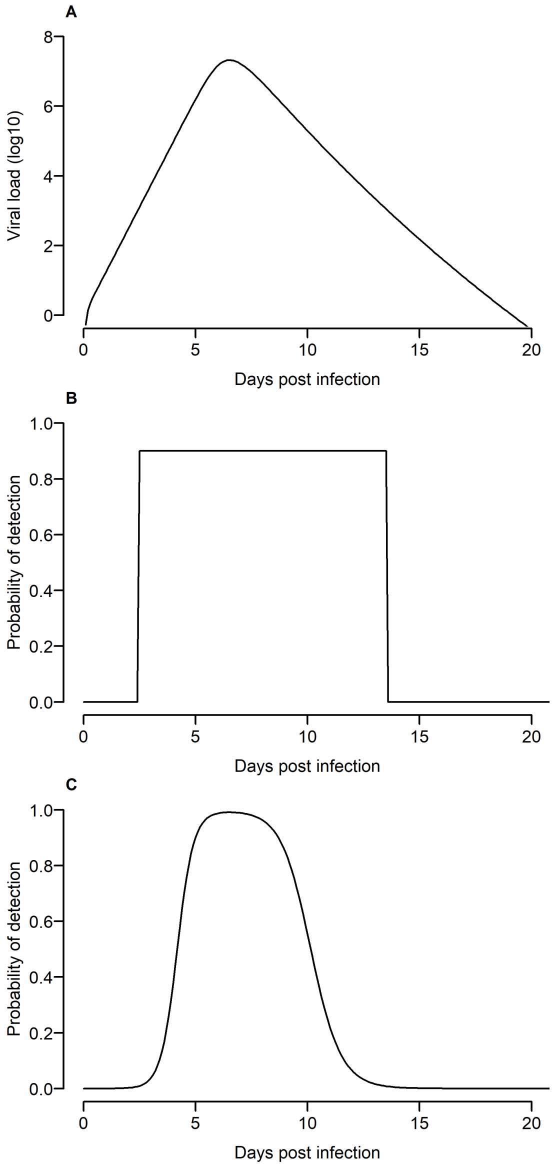

**Figure S6. Probability of infection detection for the RT-PCR test and the antigen test versus time since infection.** (A) Viral load evolution for a representative individual (patient 4 in the German dataset). (B) Probability of detection with a RT PCR test and (C) with an antigen test. These probabilities are calculated from the viral load versus time (e.g., in (A)), but the probabilities are plotted as time since infection for visualization.

#### Supplementary Tables

**Table S1. Estimated the time of infection for the individuals in the NBA dataset.** We fitted both the TCL and the innate immune model to the viral load data. The maximum likelihoods (as -2*log-likelihood, i.e. -2*LL) are reported.

| Model | -2*LL | Estimated time of infection (days) prior to the measured peak viral load | | | | | | | | |
|---|---|---|---|---|---|---|---|---|---|---|
| | | 737 | 755 | 942 | 1273 | 1740 | 2349 | 2463 | 3485 | 3491 |
| **TCL** | 227.4 | 6.9 | 6.2 | 5.6 | 6.0 | 6.2 | 9.6 | 6.5 | 9.7 | 7.1 |
| **Innate Immune** | 221.3 | 6.7 | 5.8 | 5.1 | 5.4 | 5.8 | 9.2 | 6.0 | 9.2 | 6.7 |

**Table S2. Model comparison using AIC scores.** The models considered are the target cell limited (TCL) model and three versions of the innate immunity model. The best model, i.e. the model with the lowest AIC score is the model assuming that assuming interferon signaling turns target cells into refractory cells, i.e the innate immunity – refractory cell model. For this model, we further tested if the source of dataset covaries with fitted parameters of the model. We fitted the model first assuming that all parameters covary with the covariate. We then exclude the parameter that has the lowest p-value by the Pearson's correlation test, which tests whether covariates should be removed from the model by Monolix. The models with the covariate are not significantly better than the model without the covariate, and thus there is no significant statistical evidence that the two datasets differ in their parameter values.

| Model | Parameters that covary with the source of data | -2 log-likelihood | AIC |
|---|---|---|---|
| **TCL** | none | 479.1 | 495.1 |
| **Innate Immunity – reducing infectivity** | none | 479.1 | 499.1 |
| **Innate Immunity – reducing virus production** | none | 478.1 | 498.1 |
| **Innate Immunity – refractory cells** | none | 447.1 | 472.0 |
| | $\beta, \delta, \pi, \Phi, \rho$ | 442 | 480.0 |
| | $\beta, \delta, \pi, \Phi$ | 442.2 | 478.2 |
| | $\beta, \delta, \pi$ | 442.8 | 476.8 |
| | $\beta, \delta$ | 442.9 | 472.9 |
| | $\beta$ | 443.2 | 472.2 |
| | $\delta$ | 444.6 | 472.6 |

**Table S3. Estimated individual parameter values.** The means and standard deviations (SD) are calculated in Monolix assuming that individual parameters follow log-normal distributions.

| ID* | $\beta$** ($10^{-8}$ mL/day) | $\delta$ (/day) | $\pi$ (/day) | $\Phi$** ($10^{-6}$ mL/day) | $\rho$ (/day) | $R_{0,within}$ |
|---|---|---|---|---|---|---|
| **737** | 1.58 | 1.8 | 37.5 | 0.7 | 0.004 | 2.6 |
| **755** | 4.26 | 1.4 | 50.9 | 0.2 | 0.004 | 12.7 |
| **942** | 5.37 | 1.8 | 50.9 | 2.4 | 0.004 | 12 |
| **1273** | 2.92 | 1.8 | 44.5 | 0.4 | 0.004 | 5.7 |
| **1740** | 1.83 | 2 | 36.7 | 15.6 | 0.004 | 2.7 |
| **2349** | 5.12 | 1.4 | 51.4 | 1.8 | 0.004 | 14.9 |
| **2463** | 2.94 | 1.8 | 45.8 | 0.3 | 0.004 | 5.9 |
| **3485** | 2.1 | 1.3 | 43 | 0.4 | 0.005 | 5.4 |
| **3491** | 1.83 | 1.7 | 40.3 | 0.4 | 0.004 | 3.5 |
| **1** | 5.29 | 1.8 | 50.2 | 5.9 | 0.005 | 12 |
| **2** | 3.59 | 2.1 | 45.9 | 0.6 | 0.004 | 6.2 |
| **3** | 5.16 | 1.8 | 48.9 | 20.2 | 0.004 | 11 |
| **4** | 3.94 | 2.2 | 47 | 1 | 0.004 | 6.7 |
| **7** | 3.08 | 2 | 44.5 | 0.4 | 0.004 | 5.5 |
| **8** | 3.74 | 1.6 | 46.9 | 1.5 | 0.005 | 9 |
| **10** | 1.97 | 1.3 | 40.5 | 1.5 | 0.005 | 4.7 |
| **14** | 2.76 | 1.8 | 42.3 | 5.1 | 0.004 | 5.1 |
| **Mean** | **3.38** | **1.7** | **45.1** | **3.4** | **0.004** | **7.4** |
| **SD** | **1.31** | **0.26** | **4.61** | **5.74** | **0.0001** | **3.79** |

* The first 9 IDs are according to the IDs reported in Ref. (17) and the rest of IDs are according to the IDs reported in Ref. (16).

** The unit for $\beta$ and $\Phi$ are mL/day for the NBA dataset and swab/day for the German dataset.

**Table S4. Sensitivity of estimated parameter values against variations in the values of fixed parameters in the innate immune model.**

| Model assumption* | $\boldsymbol{\beta}$** ($10^{-8}$ mL /day) | $\boldsymbol{\delta}$ (/day) | $\boldsymbol{\pi}$ (/day) | $\boldsymbol{\Phi}$** ($10^{-6}$ mL /day) | $\boldsymbol{\rho}$ (/day) | $R_{0,within}$*** | | AIC |
|---|---|---|---|---|---|---|---|---|
| | | | | | | mean | std | |
| **Baseline Model** | 3.2 | 1.7 | 45.3 | 1.3 | 0.004 | 7.4 | 3.8 | 472.1 |
| $\boldsymbol{E_0 = 5}$ **cells** | 5.3 | 1.8 | 25 | 0.4 | 0.007 | 6.1 | 2.7 | 471.4 |
| $\boldsymbol{E_0 = 10}$ **cells** | 4.5 | 1.9 | 27.7 | 0.4 | 0.007 | 5.8 | 2.4 | 472.3 |
| $\boldsymbol{c = 5/\text{day}}$ | 2.6 | 1.8 | 34.8 | 1.8 | 0.004 | 8.9 | 5 | 472.3 |
| $\boldsymbol{c = 20/\text{day}}$ | 8.9 | 1.5 | 31.4 | 0.3 | 0.003 | 8.3 | 3.8 | 470.9 |
| $\boldsymbol{k = 3/\text{day}}$ | 5.2 | 1.9 | 35.2 | 0.7 | 0.004 | 8.7 | 4.4 | 471.5 |
| $\boldsymbol{k = 6/\text{day}}$ | 4 | 1.6 | 31.7 | 0.7 | 0.003 | 7 | 3.2 | 471.8 |

* In each model, the fixed parameter values are set to the same values as in the baseline model except for the parameters stated.

** The unit for $\beta$ and $\Phi$ are mL/day for the NBA dataset and swab/day for the German dataset.

** The mean and standard deviation (std) of $R_{0,within}$ are calculated from the estimated values of $R_{0,within}$ from estimated individual parameters from each individual.

**Table S5. AIC scores of three models to the datasets from Jaafar et al. (12), Jones et al. (18) and Kohmer et al. (13)).**

| **Model** | **AIC** | | |
|---|---|---|---|
| | **Jaafar et al.** | **Jones et al.** | **Kohmer et al.** |
| ***Linear*** | 8171.1 | 371.8 | 76.2 |
| ***Power law*** | 282.0 | 298.6 | 58.8 |
| ***Saturation*** | 137.0 | 298.7 | 60.8 |

**Table S6. Best-fit parameter values of the three models, i.e. the linear model, the power-law model and the saturation model, describing data from Jaafar et al. (12), Jones et al. (18) and Kohmer et al. (13)).**

| **Model** | **Parameter** | **Best-fit parameter values and AIC** | | |
|---|---|---|---|---|
| | | **Jaafar et al.** | **Jones et al.** | **Kohmer et al.** |
| ***Linear*** | ***D*** | $3.8\times10^{-9}$ | $2.4\times10^{-9}$ | $2.6\times10^{-6}$ |
| ***Power-law*** | ***G*** | 0.018 | $2.3\times10^{-5}$ | 0.003 |
| | ***h*** | 0.24 | 0.53 | 0.45 |
| ***Saturation*** | ***J*** | 2.3 | 2.1 | 3.7 |
| | ***h*** | 0.51 | 0.69 | 0.62 |
| | ***Km*** | $8.9\times10^{6}$ copies/ml | $6.6\times10^{8}$ copies | $1.0\times10^{6}$ copies/ml |
